# Genome-wide Association Study of Social Isolation in 63 497 Japanese Individuals from the General Population

**DOI:** 10.1101/2024.09.21.24314109

**Authors:** Hisashi Ohseto, Kosuke Inoue, Ippei Takahashi, Taku Obara, Akira Narita, Mami Ishikuro, Masatsugu Orui, Keiko Murakami, Aoi Noda, Genki Shinoda, Masato Takase, Naoki Nakaya, Mana Kogure, Rieko Hatanaka, Kumi Nakaya, Ippei Chiba, Sayuri Tokioka, Yuka Kotozaki, Atsushi Shimizu, Kozo Tanno, Atsushi Hozawa, Gen Tamiya, Naoki Kondo, Shinichi Kuriyama

## Abstract

Social isolation, characterized by a lack of social connections with family, friends, and others, is associated with adverse health outcomes. However, the genetic contribution to the susceptibility to social isolation remains unclear. This study aimed to identify genetic loci associated with social isolation using the Lubben Social Network Scale (LSNS-6) in a Japanese population. The Tohoku Medical Megabank Community-Based Cohort Study was conducted between 2013 and 2016. The participants were genotyped using the Affymetrix Axiom Japonica Array. The LSNS-6 was used to assess familial and friend ties through six questions and social isolation statuses were defined as total scale, family subscale, and friend subscale. Genome-wide association studies (GWASs) were conducted using a generalized linear mixed model, adjusting for age, sex, and 10 genetic principal components. In total, 63 497 participants who completed genotyping and the LSNS-6 were included. The mean age was 59.4±11.9 years, and 41 126 (64.8%) were female. Significant genetic loci were identified in GWASs for the total scale (rs10736933 near ACADSB and HMX3) and friend subscale of LSNS-6 (rs1778366 near LINC02315 and LRFN5). This study provides the first genome-wide evidence of social isolation in the Japanese population, suggesting associations with ACADSB, HMX3, LINC02315, and LRFN5. These findings could enable personalized prevention and intervention for social isolation and related psychiatric disorders.

## Introduction

The epidemic of social isolation, a concept defined as a lack of quality and quantity in social connections with family members, friends, and others,^1^ has been an underappreciated public health crisis that has harmed individual and societal health.^2^ This condition is linked to mental health,^3,4^ cardiovascular disease,^5^ and mortality.^6^ Social factors play a significant role in social isolation, but it is important to recognize that it is not just a result of external influences. Social isolation also has a biological basis, and relevant genetic and pathological factors need to be explored.^7–9^ From a psychiatric perspective, a deficit or reduction in social interaction, which is closely related to social isolation, is often observed in patients with autism spectrum disorders,^10^ major depressive disorder,^11^ schizophrenia,^12^ and Alzheimer’s disease.^13^ A previous study revealed that these psychiatric disorders share common genetic variant risks,^14^ which may partly be explained by a common genetic predisposition for social isolation. Therefore, establishing the genetic architecture of social isolation is imperative to understand its role in psychiatric conditions and other health outcomes.

Currently, four genome-wide association studies (GWASs) of social isolation^15–18^ have been registered in the GWAS catalog^19^ (trait ID: EFO_0009592; access date: Oct 21st, 2023), all of which were conducted using the UK Biobank database. Although these studies identified more than 30 associations under various definitions of social isolation, challenges persist in this research area. First, the existing studies lack validated questionnaires for social isolation assessment. Typically, these studies utilized either singular or a combination of self-report questionnaires, which lack thorough validation in terms of consistency, structural stability, and clinical correlation. The need for validated measures is critical for reliable replication, considering the variability in the reported loci across different GWASs, which is likely due to the varying definitions of social isolation. Second, no GWAS of social isolation has been conducted among East Asian populations. The GWAS of social isolation in East Asian populations is important, especially given the distinct allele frequencies and linkage disequilibrium between East Asian and European populations. In addition, the prevalence of social isolation is higher in Japan than in the UK,^20^ accentuating the need for genetic studies on social isolation in East Asian populations.

Therefore, we conducted GWASs of social isolation, defined by a validated questionnaire—the Lubben Social Network Scale (LSNS-6)^21^—in East Asian populations using population-based cohorts of more than 60 000 people in Miyagi and Iwate Prefectures in Japan.

## Method

### Population

The Tohoku Medical Megabank (TMM) Project,^22^ which was initiated after the Great East Japan Earthquake, was conducted by the Tohoku University Tohoku Medical Megabank Organization (ToMMo) and the Iwate Medical University Iwate Tohoku Medical Megabank Organization (IMM). ToMMo and IMM conducted the TMM Community-Based Cohort Study (TMM CommCohort Study),^23^ a population-based adult cohort study using two different recruitment methods. Type 1 surveys were conducted among those who underwent a specific municipal health checkup, which was provided by insurers and covered all persons living in Japan. Type 2 surveys were conducted among those who voluntarily visited the assessment center through mass media, distribution of advertising flyers to public institutions, and advertisements at specific health checkup sites. Therefore, our study included four cohorts: type 1 survey conducted by the ToMMo, type 2 survey conducted by the ToMMo, type 1 survey conducted by the IMM, and type 2 survey conducted by the IMM. For the type 1 surveys, 97 419 people were invited, of whom 67 355 participated, and for the type 2 surveys, 19 846 participants were recruited. A total of 87 201 participants were included in this study. Participants who withdrew their consent (n = 4 704), those who did not have a genotype (n = 15 800) or outcome information (n = 4 017), and those who had >4 standard deviations for the first and second genetic principal components (n = 195) were excluded (Supplementary Figure 1).

All study participants provided written informed consent, and ethical approval was obtained from the Ethics Committee of the TMM Organization (2023-4-110).

### Genotyping, imputation, and quality control

The participants were genotyped using the Affymetrix Axiom Japonica Array (v2) in 19 batches, each consisting of 50 plates. Details about the genotyping conducted in TMM have been previously described.^24^ After batch genotyping, samples with call rates < 0.95 or exhibiting unusually high identical by descent values compared to others were excluded. Additionally, variants with Hardy–Weinberg equilibrium (HWE) P-values < 1.00 × 10^−5^, minor allele frequency < 0.01, or missing fraction > 0.01 were excluded from each batch. The genotype datasets from the 19 batches were merged to obtain a direct genotype dataset in the PLINK BED format. Subsequently, principal component analysis was performed using PLINK (version 2.0)^25^ on the direct genotype dataset after excluding 3^rd^ degree relatives and projecting them onto the remaining participants. For the imputed genotype dataset, pre-phasing was conducted using SHAPEIT2.^26^ The phased genotypes were then imputed with a cross-imputed panel of 3.5KJPNv2^27^ and 1KGP3^28^ using IMPUTE4.^29^ The cross-imputation panel for 3.5KJPNv2^27^ and 1KGP3^28^ was created by the -merge_ref_panels tool in IMPUTE2.^30^ After imputation, variants with minor allele frequencies < 0.01, missing fraction > 0.05, HWE P-value < 1.00 × 10^−5^, and imputation information scores < 0.4 were excluded. Finally, 9 040 166 variants were included in the GWASs.

### Outcomes

At the baseline survey, participants answered the LSNS-6, which evaluates familial and non-familial ties, using identical sets of three questions.^21^ The familial items include “How many relatives do you see or hear from at least once a month?,” “How many relatives do you feel at ease with that you can talk about private matters?,” and “How many relatives do you feel close to such that you can call on them for help?” Similarly, the non-familial items evaluate the same aspects but with friends instead of relatives. Each LSNS-6 item is scored from 0 to 5, contributing to a total score ranging from 0 to 30, with higher scores representing broader social networks. Social isolation is defined as a score of <12, indicating an average of two or fewer connections across the six social network dimensions assessed by the LSNS-6.^21^ The three familial LSNS-6 items form the family subscale, where limited familial ties are determined by a score of less than 6. Likewise, the three nonfamilial LSNS-6 items form the friend subscale, where limited non-familial ties are determined by a score of <6.

### Statistical analysis

Age at recruitment, sex, and social isolation status were summarized for each cohort. For each social isolation status defined by the total scale, family subscale, and friend subscale, GWASs were conducted separately in the four cohorts. GCTA (1.94.0)^31^ was used with a generalized linear mixed model adjusted for age, sex, and 10 genetic principal components. The results from the four cohorts were meta-analyzed with METAL^32^ using a sample size-weighted method. Genome-wide statistical significance was set at a conventional P-value < 5 × 10^−8^. Clumping was applied using PRSice-2 (version 2.3.5)^33^ to determine independent variants, with only significant or top-hit variants reported. These variants were annotated using ANNOVAR.^34^ The genomic inflation factors were calculated for each GWAS meta-analysis by dividing the median of the observed chi-squared test statistics by the expected median of the corresponding chi-squared distribution. Heritability was estimated for each GWAS meta-analysis by LDSC (1.0.1).^35^ For the significant or top-hit variants, the relationship between genotype and social isolation status was assessed for each cohort by logistic regression analyses adjusted for age, sex, and 10 genetic principal components, and then meta-analyzed. In addition, cross-tabulations were performed according to the genotypes of participants’ characteristics (Supplementary Methods).

Two sensitivity analyses were conducted. First, given that LSNS-6 responses can be misclassified depending on the participants’ cognitive and mental status, the associations between significant or top-hit variants and social isolation status were analyzed in a population excluding participants who self-reported dementia or major depressive disorder. Second, considering that the impact of the Great East Japan Earthquake varied geographically and affected social ties, the analysis was conducted in a population excluding participants who reported that their houses were partially or completely damaged by the earthquake.

## Results

A total of 63 497 participants were included in the ToMMo type 1 survey (n = 28 835), ToMMo type 2 survey (n = 9 510), IMM type 1 survey (n = 17 608), and IMM type 2 survey (n = 7 544) (Supplementary Figure 1). The mean age was 59.4 ± 11.9 years, and 41 126 (64.8%) participants were female. A total of 16 353 (25.8%), 9 418 (14.8%), and 21 638 (34.1%) participants showed social isolation on the total scale, family subscale, and friend subscale, respectively (Table 1). In each of the four cohorts, the number and percentage of socially isolated participants in the total scale were 7 339 (25.5%), 2 007 (21.1%), 4 940 (28.1%), and 2 067 (27.4%), respectively.

**Table 1.** Characteristics of the four cohorts in the TMM CommCohort Study

|  | ToMMo |  | IMM |  |
| --- | --- | --- | --- | --- |
|  | Type 1 | Type 2 | Type 1 | Type 2 |
|  | survey | survey | survey | survey |
| n | 28 835 | 9 510 | 17 608 | 7 544 |
| Female, % | 17 903 (62.1) | 7 249 (76.2) | 10 947 (62.2) | 5 027 (66.6) |
| Age at recruitment,<br>years | 59.62 ± 11.55 | 57.71 ± 13.36 | 61.32 ± 10.61 | 56.38 ± 13.26 |
| Social isolation status |  |  |  |  |
| Total scale, % | 7 339 (25.5) | 2 007 (21.1) | 4 940 (28.1) | 2 067 (27.4) |
| Family subscale, % | 4 212 (14.6) | 1 396 (14.7) | 2 678 (15.2) | 1 132 (15.0) |
| Friend subscale, % | 9 754 (33.8) | 2 619 (27.5) | 6 514 (37.0) | 2 751 (36.5) |

TMM CommCohort Study: Tohoku Medical Megabank Community-Based Cohort Study; ToMMo: Tohoku University Tohoku Medical Megabank Organization; IMM: Iwate Medical University; Iwate Tohoku Medical Megabank Organization. Data are shown as the mean ± standard deviation for continuous variables and n (%) for categorical variables. Social isolation was evaluated using the Lubben Social Network Scale. The total score ranged from 0 to 30. Scores < 12 were considered socially isolated. The family and friend subscales were calculated by summing up each of the three items, and the total ranged from 0 to 15. Scores < 6 were considered socially isolated.

Two loci were significant in the GWAS meta-analyses (Figure 1). The significant variant in the total scale GWAS was rs10736933, located at 10q26.13 (Table 2). The rs10736933 variant has been annotated as an intergenic variant between Short/branched chain acyl-CoA dehydrogenase (ACADSB) and H6 family homeobox 3 (HMX3). ACADSB is a member of the acyl-CoA dehydrogenase family of enzymes with activity toward the short/branched chain acyl-CoA derivatives.^36^ HMX3 is expected to have DNA-binding transcription factor activity, especially in nerve cell types.^37,38^ The significant variant in the friend subscale GWAS was rs1778366, located at 14q21.1 (Table 2). The rs1778366 was annotated as an intergenic variant between LINC02315, leucine-rich repeats, and fibronectin type III domain-containing protein 5 (LRFN5). LINC02315 is a long intergenic non-protein-coding RNA. LRFN5, also known as synaptic adhesion-like molecule 5 (SALM5), is a small transmembrane protein gene that belongs to a family of postsynaptic organizers involved in synaptic development, organization, and plasticity.^39,40^ A top-hit but not significant variant in the family subscale GWAS was rs780848113, located at 1q32.1 (Table 2), annotated as an intergenic variant between leiomodin 1 (LMOD1) and the translocase of the inner mitochondrial membrane 17A (TIMM17A). Each of the three variants was associated to a different degree with the three scales of social isolation, suggesting a scale-specific association (Figure 2). Those with the alternative allele of rs780848113, which was the top hit in the family subscale GWAS, had lower hemoglobin A1c measurements, while those with the alternative allele of rs1778366, which was significant in the friend subscale GWAS, had a higher prevalence of depressive symptoms (Supplementary Tables 1-3). Other participant characteristics were not associated with the genotypes of individual variants.

**Figure 1.**
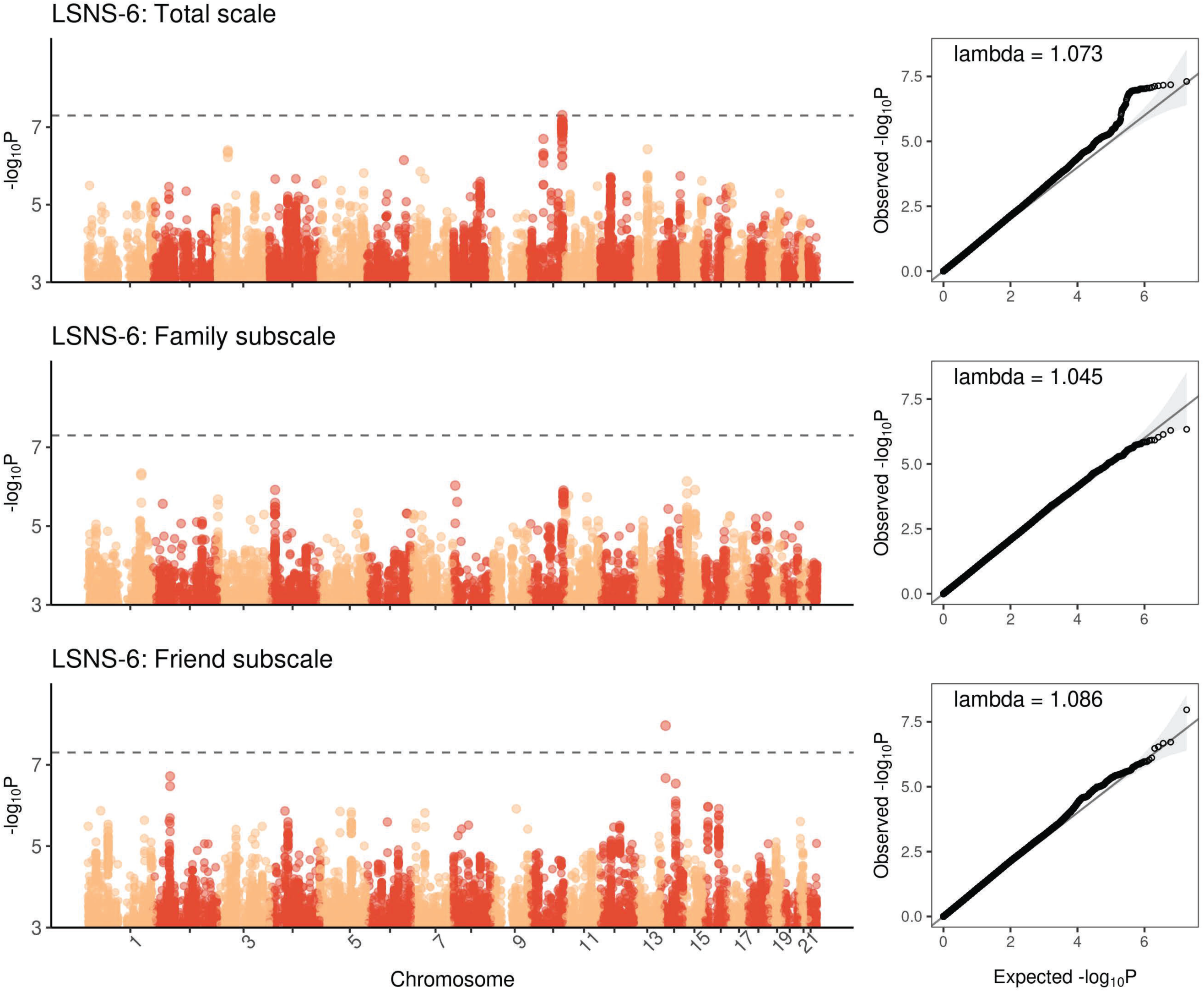
GWAS meta-analyses for social isolation status The Manhattan and QQ plots for social isolation status are presented. Three social isolation statuses—the total scale, family subscale, and friend subscale—were measured using the LSNS-6. One locus in the total scale GWAS and one in the friend subscale GWAS were significant. GWAS: Genome-wide association study; QQ plot: Quantile-quantile plot; LSNS-6: Lubben Social Network Scale.

**Table 2.**
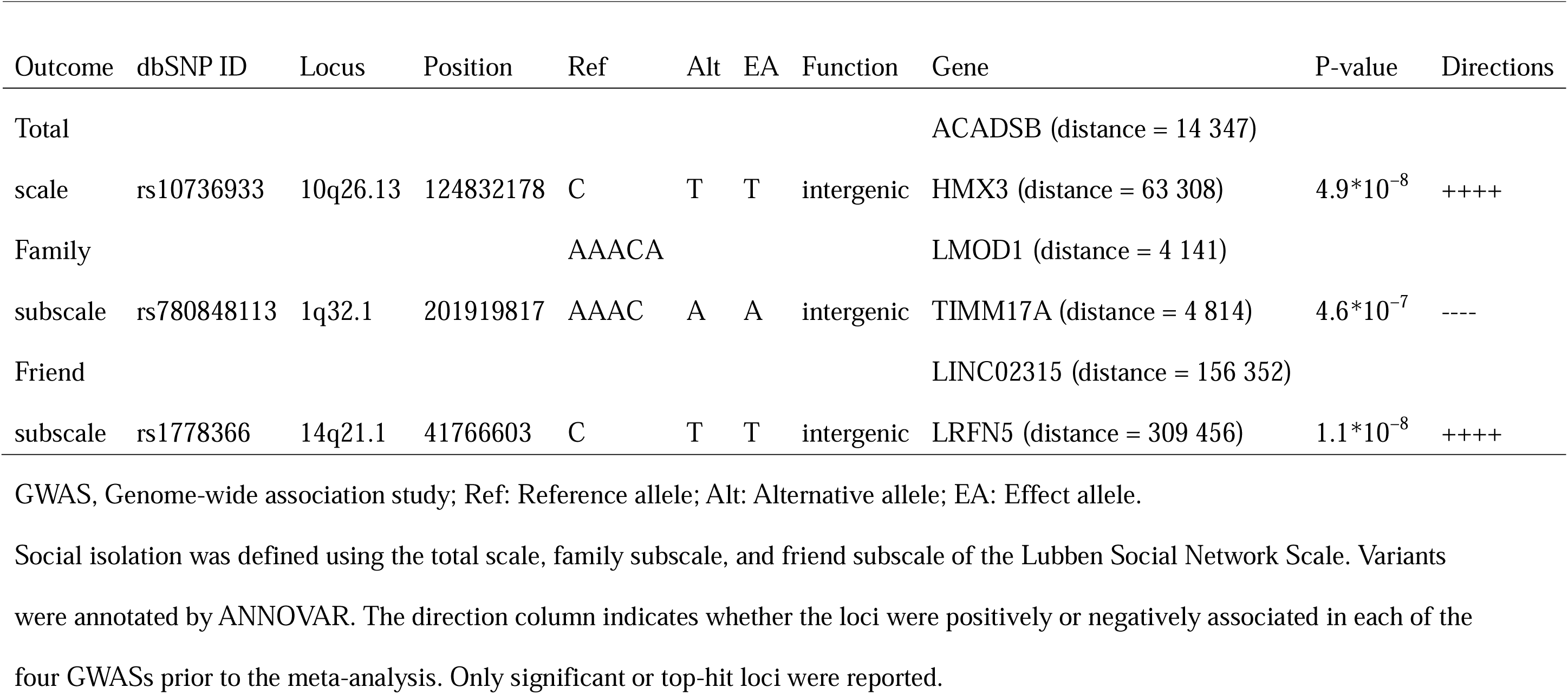
Significant variants in GWAS meta-analyses of social isolation status

| Outcome | dbSNP ID | Locus | Position | Ref | Alt | EA | Function | Gene | P-value | Directions |
| --- | --- | --- | --- | --- | --- | --- | --- | --- | --- | --- |
| Total |  |  |  |  |  |  |  | ACADSB (distance = 14 347) |  |  |
| scale | rs10736933 | 10q26.13 | 124832178 | C | T | T | intergenic | HMX3 (distance = 63 308) | $4.9 \times 10^{-8}$ | ++++ |
| Family |  |  |  | AAACA |  |  |  | LMOD1 (distance = 4 141) |  |  |
| subscale | rs780848113 | 1q32.1 | 201919817 | AAAC | A | A | intergenic | TIMM17A (distance = 4 814) | $4.6 \times 10^{-7}$ | ---- |
| Friend |  |  |  |  |  |  |  | LINC02315 (distance = 156 352) |  |  |
| subscale | rs1778366 | 14q21.1 | 41766603 | C | T | T | intergenic | LRFN5 (distance = 309 456) | $1.1 \times 10^{-8}$ | ++++ |

GWAS, Genome-wide association study; Ref: Reference allele; Alt: Alternative allele; EA: Effect allele.
Social isolation was defined using the total scale, family subscale, and friend subscale of the Lubben Social Network Scale. Variants were annotated by ANNOVAR. The direction column indicates whether the loci were positively or negatively associated in each of the four GWASs prior to the meta-analysis. Only significant or top-hit loci were reported.

**Figure 2.**
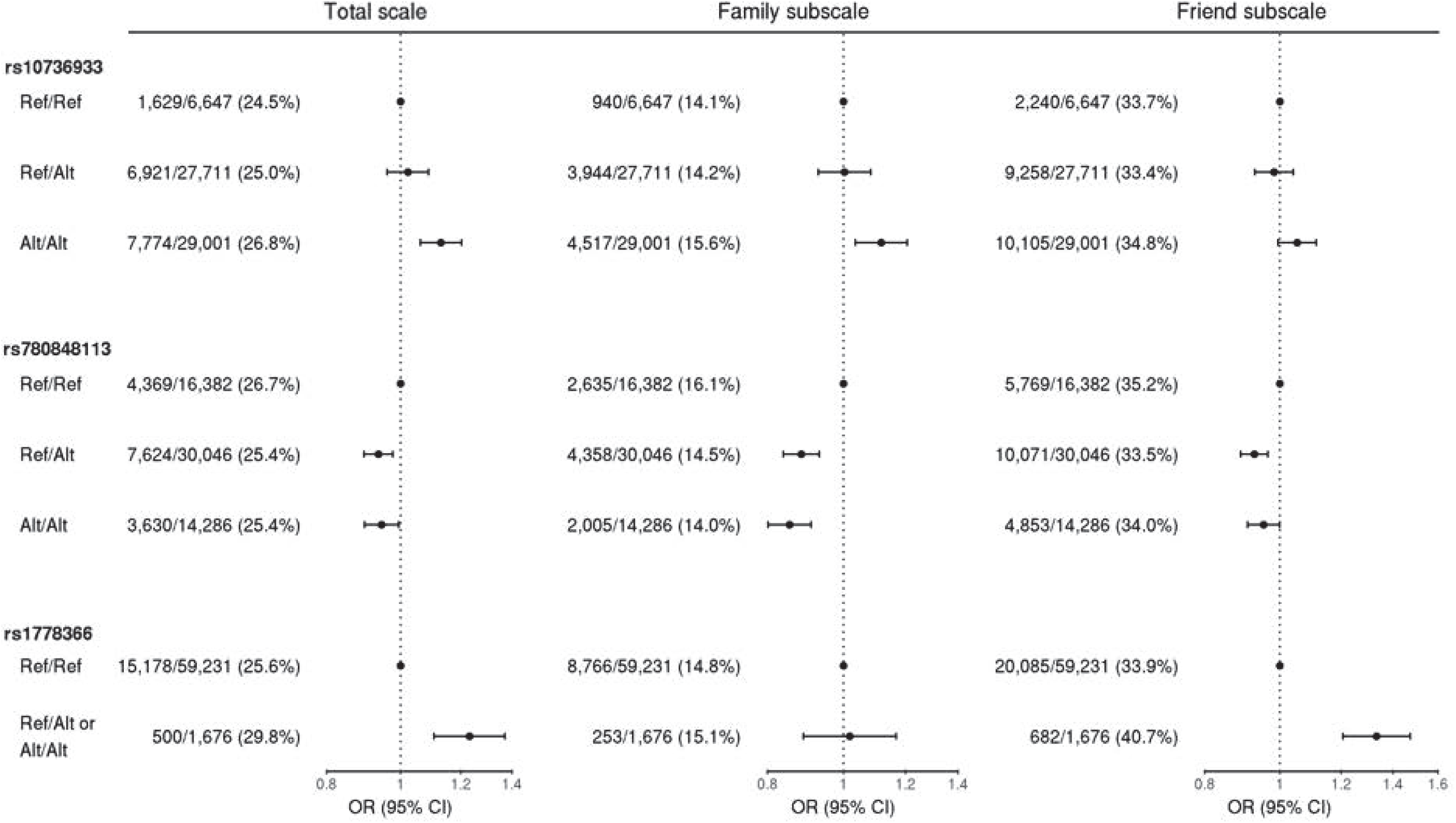
The relationship between genotype and social isolation status For significant or top-hit variants, the relationships between genotype and social isolation status were assessed for each cohort by logistic regression analyses adjusted for age, sex, and 10 genetic principal components, and then meta-analyzed. Ref: Reference allele; Alt: Alternative allele; OR: Odds ratio; CI: Confidence interval.

Both variants were positively associated with social isolation across all four cohorts (Table 3). The genomic inflation factors, lambda, were 1.073, 1.045, and 1.086 in the GWASs of the total scale, family subscale, and friend subscale, respectively (Figure 1). Heritability was estimated at 4.0%, 2.3%, and 3.8% for the total, family, and friend subscales, respectively. The sensitivity analyses showed results comparable to those of the main analysis (Supplementary Table 4).

**Table 3.**
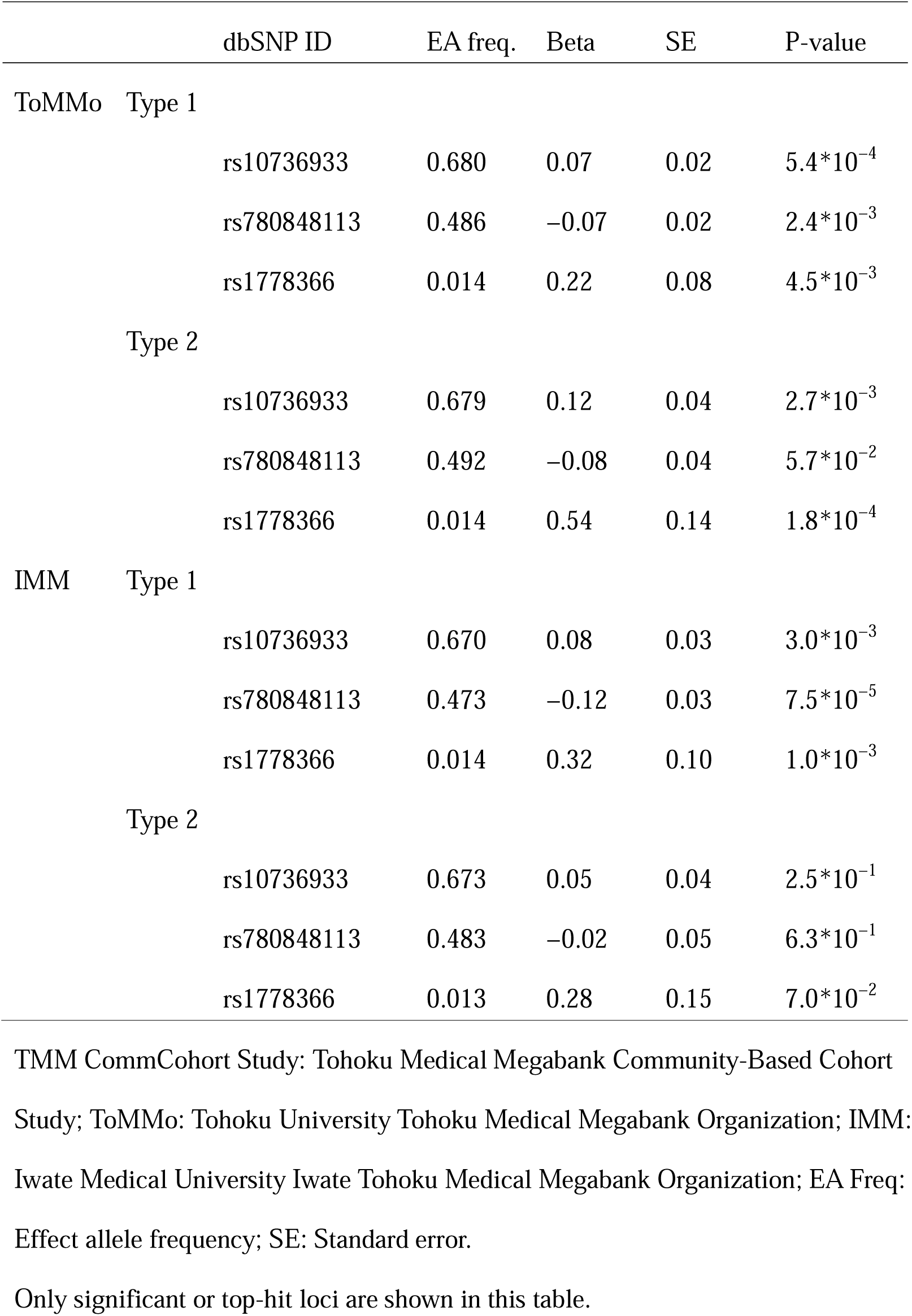
GWAS results of the four cohorts in the TMM CommCohort Study

## Discussion

Using population-based cohorts of more than 60 000 people in Japan, we identified two loci associated with social isolation: rs10736933 near ACADSB and HMX3, and rs1778366 near LINC02315 and LRFN5. These variants showed consistent associations across the four cohorts, and the heritability estimates for social isolation ranged from 2.3% to 4.0%.

Two previous GWASs have reported significant genetic variants associated with social isolation. The first study^15^ using the UK Biobank in 2018 assessed information on social isolation status based on three questions (loneliness, rarely interacting with others, and ability to confide) and identified 15 distinct genome-wide significant loci, including BPTF, GPX1, and MTCH2. The second study^16^ using the UK Biobank in 2021, assessed sociability scores based on four social functioning-related self-report questions and identified 18 independent genome-wide significant loci, including ARNTL, DRD2, and ELAVL2. Although both studies were conducted in the UK Biobank, the genome-wide significant loci were quite different, possibly because of differences in social isolation definitions. Our GWAS, conducted outside the UK Biobank, provides novel insights by identifying genes not previously associated with social isolation. This could be attributed to several factors. First, our study was the first to use a validated questionnaire, the LSNS-6, to evaluate social isolation. Second, the current study was a GWAS conducted outside the UK Biobank, which may have led to different results. Social isolation, which is a complex interplay of genetic and environmental factors, suggests that gene-environment interactions could account for the variations observed in the GWAS results. Third, differences in allele frequencies and linkage disequilibrium may alter the strength of associations for each variant.

Our study highlighted two variants, rs10736933 and rs1778366, around three protein-coding genes: ACADSB, HMX3, and LRFN5. The pattern of association between genotypes and the three scales of social isolation differed, suggesting a scale-specific etiology and genetic basis. In the GWAS catalog, intergenic variants between ACADSB and HMX3 were reported to be associated with six traits, including Lewy body dementia^41^ and response to antidepressant.^42^ As a mendelian disease gene, ACADSB was reported to cause short/branched-chain acyl-CoA dehydrogenase deficiency, a rare autosomal recessive inborn error of metabolism. This causes neurometabolic symptoms such as epilepsy and developmental delay.^43^ In GWAS catalog, variants around LRFN5 were reported in 57 traits, nearly half of which are neurobehavioral or psychiatric phenotypes, including well-being,^44^ neuroticism,^45^ educational attainment,^46^ major depressive disorder,^47^ and generalized anxiety disorder.^48^ Moreover, structural changes in LRFN5 were reported to be associated with developmental delay and autism in several cases.^49^ In this study, rs1778366 was associated with depressive symptoms, indicating a possible involvement of psychiatric etiology in social isolation in the friend subscale. In summary, our GWAS findings suggest the genetic relevance of social isolation to neurobehavioral or psychiatric phenotypes.

Two recent post-GWAS of social isolation revealed a relationship between social isolation and neurobehavioral or psychiatric phenotypes. The first study^15^ revealed a significant enrichment in the expression of social isolation-associated genes in the central nervous system and strong positive genetic overlaps with neuroticism and depressive symptoms. The second study^16^ showed that the sociability score (the higher the score, the more sociable) had negative genetic correlations with autism spectrum disorders, depression, and schizophrenia. These two studies highlighted that social isolation has its own genetic background and is genetically related to other neurobehavioral and psychiatric phenotypes. Consistent with these findings, our study highlighted the genes implicated in these conditions. When considering heritability, previous studies have estimated the heritability of social isolation at 4.2%^15^ and 6.0%,^16^ which were higher than our results: 4.0%, 2.3%, and 3.8%. This may be because the LSNS-6 focused on objective facts, such as “How many relatives do you see or hear from at least once a month?,” whereas previous studies have also used subjective loneliness to measure social isolation; thus, the LSNS-6 was less affected by the psychological state of the participants, which generally exhibits high heritability.^50^ The distinction between objective and subjective measures of social isolation is critical and warrants careful consideration in future research.

## Implications

This study has two implications. First, it reveals two novel susceptibility loci that provide insights into the biological mechanisms involved in social isolation. Because reduced social interaction is a common symptom of psychiatric disorders, these loci may provide clues to the pathogenesis of psychiatric disorders. Second, our GWAS results can be used for further post-GWAS research in East Asian populations. Methodologies such as genetic correlation, cell-type enrichment, and Mendelian randomization are potential approaches for uncovering significant insights into the impact of social isolation on health but require ethnic concordance with the target population. Given that the prevalence of social isolation is higher in Japan than in the UK,^20^ we hope that future post-GWAS will help us understand its pathology and guide effective interventions for East Asian populations.

### Strength and limitations

Our study is the first to conduct a GWAS of social isolation in East Asian populations, highlighting two novel loci. Social isolation was measured using a validated questionnaire (the LSNS-6). However, this study has several limitations. First, owing to the nature of the TMM project, which was initiated after the Great East Japan Earthquake, it is possible that some of the TMM project participants were affected by the disaster,^51–53^ which limits the generalizability of the present results. Sensitivity analyses focusing on participants whose houses were less damaged by the earthquake yielded results similar to those of the main analysis. However, the impact of the earthquake on social connections could be significant, and the results should be interpreted with caution. Second, our GWAS showed slightly high genomic inflation factors, which may indicate population stratification. However, since TMM project participants were recruited from only two prefectures in Japan and are expected to be relatively homogeneous. Additionally, since social isolation is a neurobehavioral phenotype, which tends to be polygenic,^54,55^ it is reasonable to attribute part of the high genomic inflation factor to polygenicity. Third, our study population included over 60 000 participants, which was relatively small for a GWAS, since previous studies in the UK biobank included nearly 500 000 participants. This may have limited the detection power of our study.

## Conclusions

Our GWASs revealed the first genome-wide evidence for a genetic predisposition to social isolation in the East Asian population, highlighting the possible involvement of ACADSB, HMX3, LINC02315, and LRFN5.

## Data Availability

Individual data are available upon request after the approval of the Ethical Committee and the Materials and Information Distribution Review Committee of Tohoku Medical Megabank Organization.

## Acknowledgments

The authors would like to thank all the participants who consented to participate in this study and all the staff at the Tohoku Medical Megabank Organization, Tohoku University, Iwate Tohoku Medical Megabank Organization, and Iwate Medical University.

A full list of the members of the Tohoku Medical Megabank Organization is available at https://www.megabank.tohoku.ac.jp/english/a230901/.

We used the artificial intelligence tool GPT-4 language model, version 4.0, developed by OpenAI, to assist in the initial drafting and editing of this manuscript. It provided suggestions for sentence structure and grammatical corrections. However, all intellectual contributions and final edits were made by the authors.

## Authors’ contributions

Conceptualization: Hisashi Ohseto, Kosuke Inoue, and Ippei Takahashi

Methodology: Hisashi Ohseto and Ippei Takahashi

Visualization: Hisashi Ohseto

Supervision: Shinichi Kuriyama and Naoki Kondo

Writing – original draft: Hisashi Ohseto, Kosuke Inoue, and Ippei Takahashi

Writing – review & editing: All the authors

## Information on previous presentation

This work has not been previously presented anywhere.

