## Supplementary Information for "Genome-wide Association Study of Social Isolation in 63 497 Japanese Individuals from the General Population"


| Items | | Pages |
|---|---|---|
| Supplementary Method | | 2 |
| References | | 3 |
| Supplementary Tables | | |
| | Supplementary Table 1. Participant characteristics by rs10736933 genotype | 4 |
| | Supplementary Table 2. Participant characteristics by rs780848113 genotype | 5 |
| | Supplementary Table 3. Participant characteristics by rs1778366 genotype | 6 |
| | Supplementary Table 4. Sensitivity analyses for the relationship between genotype and social isolation status | 7 |
| Supplementary Figure and Supplementary Figure Legend | | |
| | Supplementary Figure 1. Flowchart of study population selection in the TMM Community-Based Cohort Study | 8 |

### Supplementary Method

Cross-tabulations were performed according to the genotypes for depressive symptoms, hemoglobin A1c measurement, non-high-density lipoprotein cholesterol measurement, blood pressure measurement, body mass index, medical history, education, and family size. Depressive symptoms were measured using the 20-item Center for Epidemiologic Studies– Depression Scale, with a score of ≥17 defined as having depressive symptoms.[1] Non-high-density lipoprotein cholesterol measurement, hemoglobin A1c measurement, blood pressure measurement, and body mass index were measured by either the specific municipal health check-up or the Tohoku Medical Megabank Project protocol.[2] Medical history (type 2 diabetes mellitus, dyslipidemia, chronic hypertension, major depressive disorder, and dementia), education, and family size were self-reported in the baseline survey. The genotypes were classified as reference/reference, reference/alternative, or alternative/alternative. For rs1778366, the frequency of the alternative allele was <0.05; thus, participants homozygous for the alternative allele were included with heterozygous participants in cross-tabulation and regression analyses.

**Supplementary Table 1. Participant characteristics by rs10736933 genotype**

| | Genotype of rs10736933 | | | |
|---|---|---|---|---|
| | **Ref/Ref** | **Ref/Alt** | **Alt/Alt** | **P-value** |
| N | 6 647 | 27 711 | 29 001 | |
| Female, % | 4 345 (65.4) | 17 934 (64.7) | 18 757 (64.7) | 0.553 |
| Age at recruitment, years | 59.38 ± 11.98 | 59.38 ± 11.93 | 59.47 ± 11.89 | 0.624 |
| Depressive symptoms, % | 1 456 (22.7) | 5 832 (21.8) | 6 332 (22.6) | 0.067 |
| Hemoglobin A1c, % | 5.62 ± 0.57 | 5.63 ± 0.58 | 5.63 ± 0.59 | 0.304 |
| Non-HDL, mg/dL | 142.94 ± 33.51 | 143.32 ± 34.07 | 143.43 ± 34.64 | 0.716 |
| Systolic blood pressure, mmHg | 126.58 ± 17.43 | 126.67 ± 17.79 | 126.81 ± 17.69 | 0.530 |
| Diastolic blood pressure, mmHg | 75.44 ± 10.86 | 75.53 ± 10.98 | 75.64 ± 10.85 | 0.268 |
| Body mass index, $kg/m^2$ | 23.31 ± 3.54 | 23.31 ± 3.57 | 23.33 ± 3.53 | 0.867 |
| Medical history | | | | |
| Type 2 diabetes mellitus, % | 284 (4.3) | 1 287 (4.7) | 1 301 (4.6) | 0.379 |
| Dyslipidemia, % | 818 (12.5) | 3 367 (12.3) | 3 480 (12.2) | 0.730 |
| Chronic hypertension, % | 1 665 (25.3) | 7 084 (25.8) | 7 353 (25.6) | 0.682 |
| Major depressive disorder, % | 184 (2.8) | 819 (3.0) | 896 (3.1) | 0.326 |
| Dementia | 14 (0.2) | 48 (0.2) | 53 (0.2) | 0.809 |
| Education | | | | 0.075 |
| High school or lower | 4 405 (67.1) | 18 260 (66.7) | 19 084 (66.6) | |
| Junior or vocational college | 1 510 (23.0) | 6 358 (23.2) | 6 829 (23.8) | |
| University or higher | 653 (9.9) | 2 770 (10.1) | 2 725 (9.5) | |
| Family size | | | | 0.325 |
| Living alone | 525 (8.0) | 2 118 (7.8) | 2 270 (8.0) | |
| Living with one person | 2 199 (33.6) | 8 994 (33.0) | 9 591 (33.6) | |
| Living with two or more people | 3 817 (58.4) | 16 153 (59.2) | 16 653 (58.4) | |

Ref: Reference allele; Alt: Alternative allele; HDL: High-density lipoprotein.

Data are shown as the mean ± standard deviation for continuous variables and n (%) for categorical variables.

### Supplementary Table 2. Participant characteristics by rs780848113 genotype

| | Genotype of rs780848113 | | | |
|---|---|---|---|---|
| | Ref/Ref | Ref/Alt | Alt/Alt | P-value |
| n | 16 382 | 30 046 | 14 286 | |
| Female, % | 10 545 (64.4) | 19 515 (65.0) | 9 301 (65.1) | 0.334 |
| Age at recruitment, years | 59.58 ± 11.88 | 59.30 ± 11.91 | 59.51 ± 11.95 | 0.032 |
| Depressive symptoms, % | 3 502 (22.2) | 6 457 (22.2) | 3 086 (22.5) | 0.819 |
| Hemoglobin A1c, % | 5.64 ± 0.59 | 5.63 ± 0.59 | 5.62 ± 0.57 | 0.044 |
| Non-HDL cholesterol, mg/dL | 142.92 ± 34.32 | 143.66 ± 34.47 | 143.31 ± 34.00 | 0.215 |
| Systolic blood pressure, mmHg | 126.82 ± 17.79 | 126.71 ± 17.67 | 126.66 ± 17.71 | 0.705 |
| Diastolic blood pressure, mmHg | 75.62 ± 10.97 | 75.55 ± 10.92 | 75.58 ± 10.82 | 0.804 |
| Body mass index, kg/m$^2$ | 23.32 ± 3.53 | 23.32 ± 3.56 | 23.30 ± 3.52 | 0.907 |
| Medical history | | | | |
| Type 2 diabetes mellitus, % | 776 (4.8) | 1 310 (4.4) | 646 (4.6) | 0.179 |
| Dyslipidemia, % | 1 980 (12.3) | 3 607 (12.2) | 1 738 (12.4) | 0.844 |
| Chronic hypertension, % | 4 164 (25.6) | 7 609 (25.5) | 3 653 (25.8) | 0.858 |
| Major depressive disorder, % | 489 (3.0) | 927 (3.1) | 410 (2.9) | 0.473 |
| Dementia | 33 (0.2) | 55 (0.2) | 22 (0.2) | 0.627 |
| Education | | | | 0.354 |
| High school or lower | 10 832 (67.1) | 19 700 (66.3) | 9 476 (67.0) | |
| Junior or vocational college | 3 723 (23.1) | 7 068 (23.8) | 3 298 (23.3) | |
| University or higher | 1 595 (9.9) | 2 925 (9.9) | 1 362 (9.6) | |
| Family size | | | | 0.705 |
| Living alone | 1 257 (7.8) | 2 338 (7.9) | 1 116 (7.9) | |
| Living with one person | 5 430 (33.7) | 9 773 (33.1) | 4 707 (33.5) | |
| Living with two or more people | 9 440 (58.5) | 17 443 (59.0) | 8 221 (58.5) | |

Ref: Reference allele; Alt: Alternative allele; HDL: High-density lipoprotein.

Data are shown as the mean ± standard deviation for continuous variables and n (%) for categorical variables.

### Supplementary Table 3. Participant characteristics by rs1778366 genotype

| | Genotype of rs1778366 | | |
|---|---|---|---|
| | Ref/Ref | Ref/Alt or Alt/Alt | P-value |
| n | 59 231 | 1 676 | |
| Female, % | 38 286 (64.6) | 1 081 (64.5) | 0.927 |
| Age at recruitment, years | 59.44 ± 11.90 | 58.84 ± 12.42 | 0.043 |
| Depressive symptoms, % | 12 678 (22.2) | 394 (24.5) | 0.027 |
| Hemoglobin A1c, % | 5.63 ± 0.58 | 5.62 ± 0.57 | 0.532 |
| Non-HDL cholesterol, mg/dL | 143.37 ± 34.28 | 143.11 ± 34.79 | 0.809 |
| Systolic blood pressure, mmHg | 126.76 ± 17.72 | 126.23 ± 17.70 | 0.226 |
| Diastolic blood pressure, mmHg | 75.58 ± 10.91 | 75.73 ± 10.65 | 0.579 |
| Body mass index, kg/m$^2$ | 23.32 ± 3.55 | 23.18 ± 3.56 | 0.109 |
| Medical history | | | |
| Type 2 diabetes mellitus, % | 2 681 (4.6) | 69 (4.2) | 0.448 |
| Dyslipidemia, % | 7 134 (12.2) | 204 (12.4) | 0.908 |
| Chronic hypertension, % | 15 089 (25.7) | 409 (24.6) | 0.331 |
| Major depressive disorder, % | 1 777 (3.1) | 52 (3.1) | 0.887 |
| Dementia | 107 (0.2) | 3 (0.2) | >0.999 |
| Education | | | 0.737 |
| High school or lower | 39 029 (66.7) | 1 118 (67.3) | |
| Junior or vocational college | 13 731 (23.5) | 388 (23.4) | |
| University or higher | 5 752 (9.8) | 154 (9.3) | |
| Family size | | | 0.630 |
| Living alone | 4 589 (7.9) | 140 (8.5) | |
| Living with one person | 19 440 (33.4) | 541 (32.9) | |
| Living with two or more people | 34 227 (58.8) | 965 (58.6) | |

Ref: Reference allele; Alt: Alternative allele; HDL: High-density lipoprotein.

Homozygous participants for the alternative alleles were included with heterozygous participants.

Data are shown as the mean ± standard deviation for continuous variables and n (%) for categorical variables.

**Supplementary Table 4. Sensitivity analyses for the relationship between genotype and social isolation status**

| Outcome | dbSNP ID | Genotype | Main analysis OR (95% CI) | Sensitivity analysis 1 OR (95% CI) | Sensitivity analysis 2 OR (95% CI) |
|---|---|---|---|---|---|
| Total scale | rs10736933 | Ref/Ref | Reference | Reference | Reference |
| | | Ref/Alt | 1.02 (0.96 to 1.09) | 1.02 (0.96 to 1.09) | 1.02 (0.95 to 1.10) |
| | | Alt/Alt | 1.13 (1.06 to 1.20) | 1.12 (1.05 to 1.20) | 1.12 (1.05 to 1.20) |
| Family subscale | rs780848113 | Ref/Ref | Reference | Reference | Reference |
| | | Ref/Alt | 0.88 (0.84 to 0.93) | 0.88 (0.84 to 0.93) | 0.88 (0.83 to 0.94) |
| | | Alt/Alt | 0.85 (0.80 to 0.91) | 0.86 (0.80 to 0.91) | 0.85 (0.79 to 0.91) |
| Friend subscale | rs1778366 | Ref/Ref | Reference | Reference | Reference |
| | | Ref/Alt or Alt/Alt | 1.33 (1.21 to 1.47) | 1.33 (1.20 to 1.47) | 1.33 (1.19 to 1.48) |

OR: Odds ratio; CI: Confidence interval; Ref: Reference allele; Alt: Alternative allele.

Sensitivity analysis 1 was conducted in a population excluding participants who self-reported dementia or major depression. Sensitivity analysis 2 was conducted in a population excluding participants who reported that their houses were partially or completely damaged by the earthquake.

ORs and 95% CIs were calculated using logistic regression analyses adjusted for age, sex, and 10 genetic principal components.

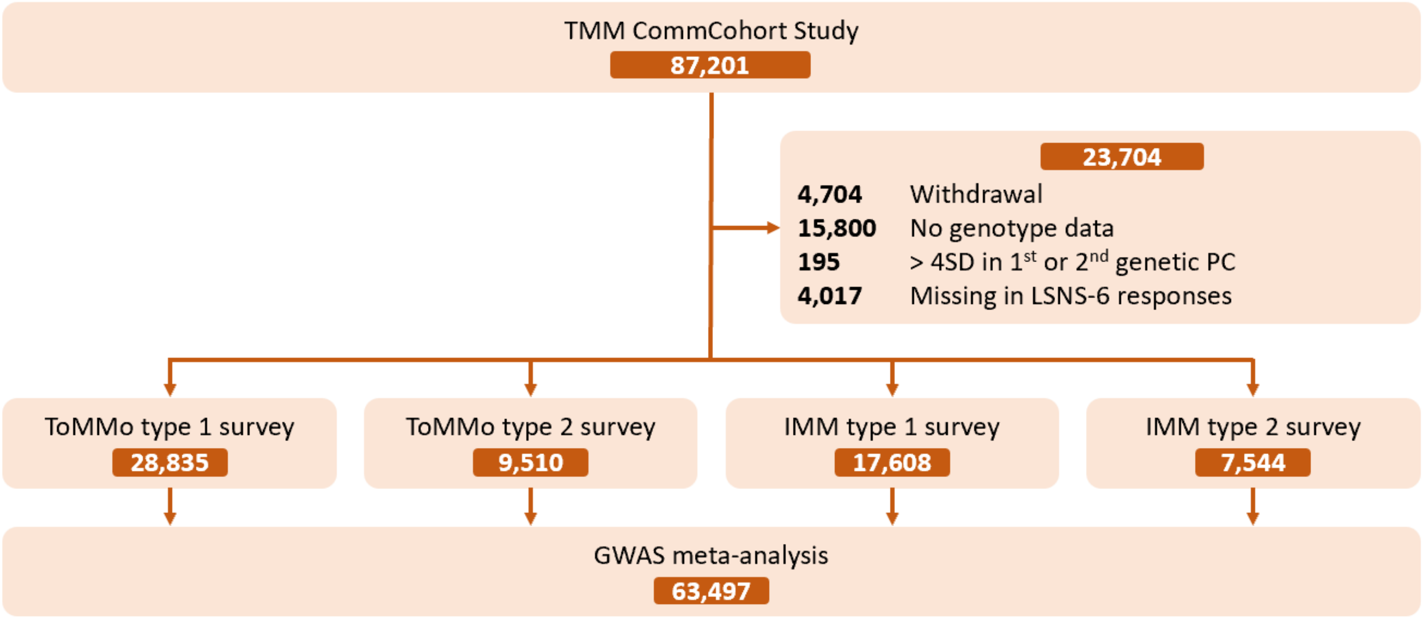


**Supplementary Figure 1. Flowchart of study population selection in the TMM Community-Based Cohort Study**

This study was conducted in four TMM community-based cohorts. All four cohorts were meta-analyzed, with a total of 63 497 participants.

TMM CommCohort Study: Tohoku Medical Megabank Community-Based Cohort Study; ToMMo: Tohoku University Tohoku Medical Megabank Organization; IMM: Iwate Medical University Iwate Tohoku Medical Megabank Organization; SD: Standard deviation; PC: Principal components; LSNS-6: Lubben Social Network Scale; GWAS: Genome-wide Association Study.
